# COVID-19 symptoms are associated with higher variability in generalized anxiety and depression: results from a one-year longitudinal study in Spain

**DOI:** 10.1101/2025.10.19.25338321

**Authors:** Marco Viola, Manolis Kogevinas, Gemma Castaño-Vinyals, Rafael de Cid, Rosalba Rosato, Guillaume Chevance

## Abstract

**Background:** The mental health impacts of COVID-19, particularly regarding anxiety and depression, are well-documented and more prevalent among women. This study aimed to assess the longitudinal effects of COVID-19 on anxiety, depression, and sleep conditions in a sample of Spanish adults.

**Methods:** A total of 608 individuals completed biweekly anxiety, depression, and sleep questionnaires for over a year (July 2021-March 2023). Data on SARS-CoV-2 infection prior to questionnaire administration were also available. Respondents were classified into three groups based on prior SARS-CoV-2 infection and symptoms during a period preceding the administration of the questionnaire (no infection, infected asymptomatic, symptomatic). Means and variability scores for anxiety, depression, and sleep scores were tracked over time for each individual. Differences between the three groups were analysed, using generalized linear models to assess the impact of COVID-19 symptoms.

**Results:** No significant differences in mean anxiety, depression, or sleep quality scores were observed among participants with no COVID-19 diagnosis, symptomatic infection, or asymptomatic infection. Women reported significantly higher anxiety and depression scores and poorer sleep quality than men (p < 0.001). Greater variability in anxiety and depression scores was found among symptomatic COVID-19 cases compared to non-infected individuals, while time since infection did not significantly influence mean scores or variability in any outcome.

**Conclusions:** Individuals with COVID-19 symptoms exhibited greater variability in anxiety and depression scores, indicating increased psychological instability. Sleep quality was not significantly affected by COVID-19 symptoms. These findings underscore the importance of targeted interventions to address post-pandemic mental health challenges.

## Introduction

The COVID-19 pandemic represents a prolonged global emergency, characterized by varying impacts across age groups, socioeconomic classes, and geographic regions. The ramifications of the pandemic extend beyond physical health, significantly increasing the vulnerability of mental health symptoms [1–2]. Existing research has documented the short and medium-term complications of COVID-19 symptoms, particularly their impact on mental health, including depression, anxiety, and sleep disorders [3–5]. Research conducted after the first COVID-19 wave revealed an alarmingly high prevalence of anxiety and depression (68.7%) among COVID-19 survivors [6].

Growing evidence highlights disparities in depression, anxiety, and sleep disorders between individuals who have contracted COVID-19 and those who have not. A study by Xie et al. found that individuals with a history of SARS-CoV-2 infection had a significant increased risk of developing anxiety and depression at one year after infection, compared to those not infected [7]. This cohort study also indicated that the odds of experiencing these mental health issues were significantly higher in individuals who had contracted the virus compared to a contemporary uninfected group [7]. Several studies and meta-analyses indicate that individuals exhibiting COVID-19 symptoms demonstrate significantly higher rates of mental health symptoms in the aftermath of infection compared to uninfected individuals [8–9]. The prevalence of depression and anxiety disorders in post-COVID-19 patients in Greece was estimated at 17.3% and 17.2%, respectively [10], which is higher than pre-pandemic levels (2.9% and 4.1%, respectively) [11]. Furthermore, it has been observed that not only mental health outcomes differ between individuals infected and uninfected by SARS-CoV-2, but also that individuals with symptomatic infection exhibited higher levels of depression and anxiety compared to those with asymptomatic infection [12].

Mental health challenges are often accompanied by physical symptoms, with sleep disturbances being among the most common. Indeed, the interplay between sleep disturbances and mental health is critical; insufficient sleep can exacerbate symptoms of anxiety and depression, while conversely, anxiety and depression can worsen sleep conditions. This can create a vicious cycle that perpetuates itself and aggravates mental health issues [13–15]. Studies suggest that people that contracted COVID-19 report insomnia at much higher rates compared to uninfected population, with an estimated prevalence of insomnia disorder among COVID-19 survivors of about 40% [16].

Most studies examining the impact of SARS-CoV-2 infection on mental health and sleep have relied on cross-sectional designs, which provide valuable insights but are limited in capturing changes over time [12,17–21]. Monitoring the long-term impact of COVID-19 is essential to identify symptom trajectories that allow the observation of their progression over time. A longitudinal study assessing trajectories of anxiety, depression, and sleep quality over time revealed that anxiety symptoms gradually declined, whereas depressive symptoms followed a more stable course [22]. This suggests that while some individuals may experience improvement, others may continue to face persistent mental health challenges long after the acute phase of infection [22].

In addition to assessing average symptom levels, it is important to examine mental health instability, a common feature of various mental health disorders, including anxiety and depression. This instability, characterized by rapid fluctuations in mood, can significantly impact clinical outcomes [23]. Specifically, instability in mental health symptoms has been associated with poorer treatment adherence, increased symptom severity, heightened risk of relapse, and diminished overall functioning in daily life [24]. This highlights that variations in mental health may reflect underlying psychological challenges that demand attention beyond average symptom levels.

This study applies a longitudinal approach to compare three distinct groups of participants: individuals without a history of SARS-CoV-2 infection, those who were infected but remained asymptomatic, and those who experienced symptomatic infection. The study focuses on differences in mean and variability levels of anxiety, depression, and sleep. Distinguishing between participants with symptoms and those without is crucial for understanding how the presence or absence of COVID-19 symptoms, rather than infection status alone, can influence mental health outcomes. We hypothesize that the presence of COVID-19 symptoms leads to a significant increase in the mean levels and variability of anxiety, depression, and sleep disturbances compared to those who are asymptomatic or have not been infected.

## Methods

### Study design and participants

This is a longitudinal study where participants were asked to complete an online questionnaire in Spanish or Catalan every two weeks for over a year, from late July 2021 to March 2023. The questionnaire assessed generalized anxiety, depression, and sleep quality. Data were collected using a GDPR-compliant smartphone application (Ethica Data). The Parc de Salut Mar Ethics Committee granted ethical approval for the study (CEIm-PS MAR, number 2020/9307/I). All participants provided written informed consent. The study population consists of a subsample of the larger GCAT epidemiological study [25] and includes an evaluation of COVID-19 within this cohort as part of the COVICAT study in Catalonia [5,26]. In the COVICAT study, 5.0% of participants were classified as COVID-19 cases. Compared to non-cases, affected individuals were slightly younger and included a marginally higher proportion of women [26]. The data retrieved from the GCAT and COVICAT studies between April and June 2023 were used exclusively for statistical analyses. These data were obtained through linkage using a unique identification code assigned to each participant, without the use of any personal information that could identify individual participants. In this longitudinal study, out of an initial sample of 873 participants, the final sample consists of 608 individuals who completed the questionnaire at least three times over time, allowing for the assessment of variability in mental health and sleep.

### Measures

#### Generalized anxiety

Anxiety was measured with the Generalized Anxiety Disorder 2-Item (GAD-2) scale, that consists of 2 items: *Over the last 2 weeks, how often have you been bothered by the following problems? 1) Feeling nervous, anxious or on edge, and 2) Not being able to stop or control worrying.* Each response has a score ranging from 0 to 3 (0: not at all; 1: several days; 2: more than half the days; 3: nearly every day), and the total score is the sum of the two scores with a total range from 0 to 6 (higher scores indicate higher anxiety). In the Spanish population, the GAD-2 demonstrates high internal consistency as well as good sensitivity and specificity, along with concurrent validity when compared to other validated measures of anxiety [27–28].

#### Depressed mood and Anhedonia

Symptoms of depression were measured at each time point using the Patient Health Questionnaire-2 (PHQ-2), a brief self-report measure of symptoms of major depressive disorder, in particular anhedonia. The PHQ-2 contains 2 items and assesses depression symptoms using the following questions: *Over the last 2 weeks, how often have you been bothered by the following problems? 1) Little interest or pleasure in doing things, and 2) Feeling down, depressed, or hopeless.* Each question has a score between 0 and 3 (0: not at all; 1: several days; 2: more than half the days; 3: nearly every day), and the total score is the sum of the two scores with a range from 0 to 6 (higher scores indicate higher depression). The PHQ-2 scale has been demonstrated to be an effective tool for the routine monitoring and assessment of symptoms related to depression in clinical settings, as evidenced by the findings of Staples et al. [29].

#### Sleep quality

Participants completed a visual analogue scale (VAS), designed to measure the overall quality of their sleep over the past two weeks. The scale ranges from 0 (“terrible”) to 10 (“excellent”). This single item measure has been validated against longer sleep questionnaires [30].

#### COVID-19 symptoms

Data on SARS-CoV-2 infection and its severity were obtained from COVID-19 positive test results or self-reported diagnoses and symptoms [5,26]. This was used to classify participants into three distinct groups: those without SARS-CoV-2 infection, asymptomatic COVID-19 individuals, and symptomatic individuals.

#### Covariates

Baseline characteristics of the participants, including age (categorized as 44–50, 51–60, and 61–71 years), gender, and educational level (classified as up to primary, secondary, and university), were recorded before the start of the longitudinal survey using pre-pandemic cohort registers [5,26]. COVID-19 vaccination status (yes/no) was also recorded, allowing the sample to be divided into two groups: vaccinated and unvaccinated individuals [31]. Finally, we also controlled for both the number of observations over time for each participant and the time, in months, that had passed since SARS-CoV-2 infection until the central observation date, calculated as the median date for each participant, with the aim of better account for the role of time on mental health outcomes.

### Statistical analyses

We used the mean levels and variability (standard deviation) of anxiety, depression, and sleep scores as the main dependent variables in this study, calculated using all available measurement occasions within individuals. First, differences in anxiety, depression, and sleep scores were examined between participants in the three groups: without SARS-CoV-2 infection, asymptomatic, and symptomatic. Second, to further evaluate the relationship between anxiety, depression, sleep, and COVID-19 symptoms, multiple linear regressions were applied. In the first model, the mean levels of anxiety, depression, and sleep scores were used as dependent variables, while group category based on COVID-19 symptoms was the independent variable. The models were adjusted for age, gender, educational level, COVID-19 vaccination status, and the number of observations per individual. Similarly, the variability of anxiety, depression, and sleep scores was analysed using the standard deviations as dependent variables. All analyses were performed with the statistical software RStudio (2023.06.01+524 release).

## Results

Table 1 presents participants demographics and COVID-19 characteristics at baseline, overall and divided by group. The average age of the sample was 55.3 years (range 44-71) with 40.5% being male and 59.5% female. The percentage of vaccinated individuals was 92.1%, and 13.3% had a symptomatic COVID-19 disease at baseline before starting the completion of the mental health questionnaire. Age, gender, and educational level exhibited no statistically significant differences among the three groups.

**Table 1.** Demographic and COVID-19 characteristics of the participants.

| | Overall | Groups | | | p (T or $\chi^2$)<sup>a</sup> |
| --- | --- | --- | --- | --- | --- |
|  |  | No COVID-19 | Asymptomatic | Symptomatic |  |
| N | 608 | 447 (73.5%) | 80 (13.2%) | 81 (13.3%) |  |
| Age, mean (S.D.) | 55.3 (7.3) | 55.5 (7.3) | 54.9 (7.9) | 54.2 (6.6) | 0.310 |
| Age range |  |  |  |  |  |
| 44-50 | 194 (31.9%) | 135 (30.2%) | 30 (37.5%) | 29 (35.8%) |  |
| 51-60 | 260 (42.8%) | 190 (42.5%) | 32 (40.0%) | 38 (46.9%) |  |
| 61-71 | 154 (25.3%) | 122 (27.3%) | 18 (22.5%) | 14 (17.3%) |  |
| Gender - female | 362 (59.5%) | 257 (57.5%) | 54 (67.5%) | 51 (63.0%) | 0.195 |
| Education level |  |  |  |  | 0.874 |
| Up to Primary school | 53 (8.7%) | 37 (8.3%) | 8 (10.0%) | 8 (9.9%) |  |
| Secondary school | 252 (41.4%) | 182 (40.7%) | 36 (45.0%) | 34 (42.0%) |  |
| University | 303 (49.8%) | 228 (51.0%) | 36 (45.0%) | 39 (48.1%) |  |
| COVID-19 vaccination - yes | 560 (92.1%) | 429 (96.0%) | 72 (90.0%) | 59 (72.8%) | < 0.001 |

<sup>a</sup>F-test for age comparison, $\chi^2$ for gender, education level and COVID-19 vaccination comparison in the three groups

The total number of analysed observations was 8310, with an average of 14 observations per individual (range: 3-26). Across all participants, the first observation was recorded in July 2021, and the last in March 2023. Figure 1 presents an example of the observed scores for anxiety, depression, and sleep for three participants, each with a different number of observations and varying symptom fluctuations over time.

**Fig 1.**
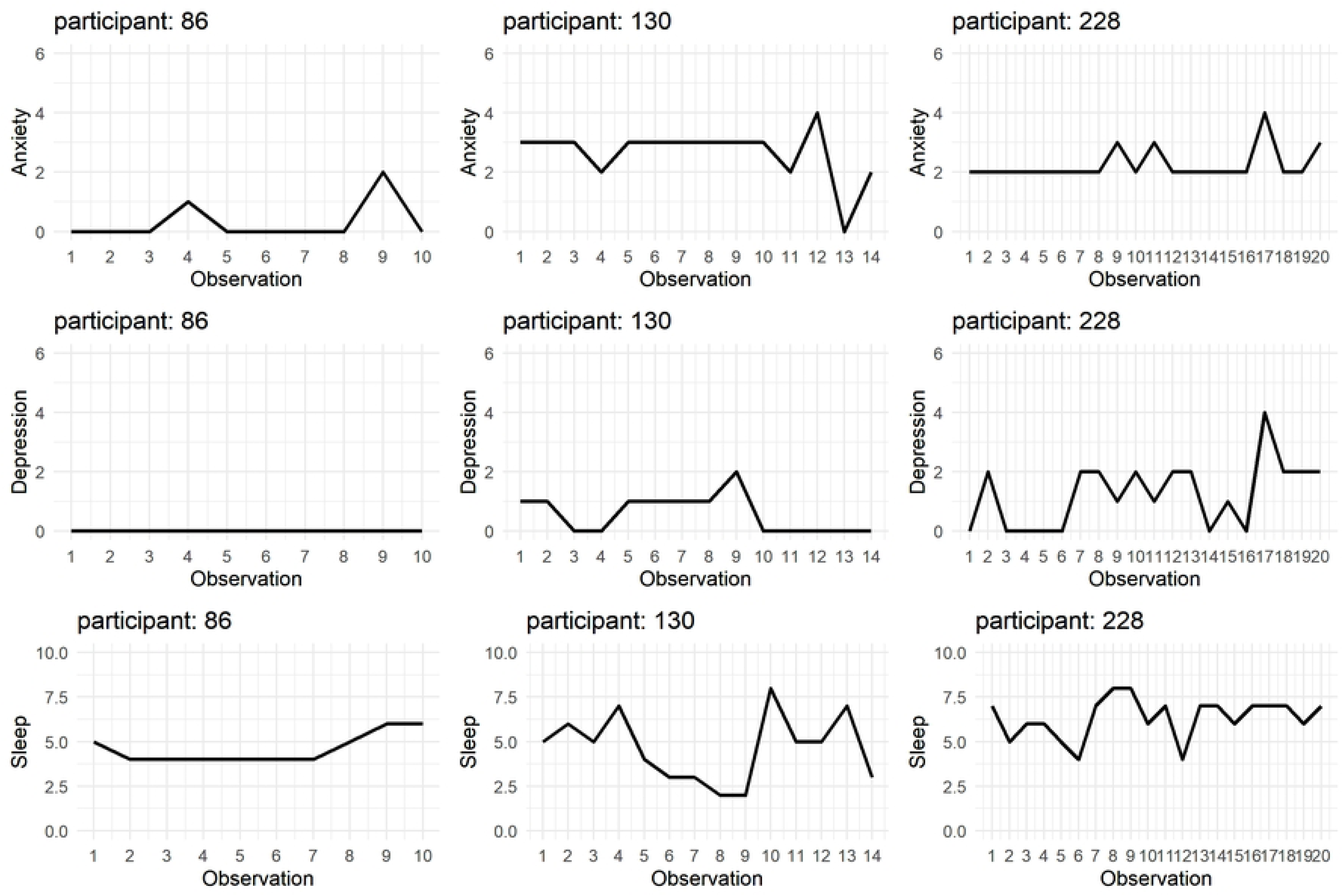
Illustration of the three study outcomes for 3 participants For participant 86, the number of observations is 10, with the observation period spanning from July 26, 2021, to January 24, 2022. For participant 130, the number of observations is 14, with the observation period spanning from July 26, 2021, to May 16, 2022. For participant 228, the number of observations is 20, with the observation period spanning from July 30, 2021, to July 15, 2022.

### Analysis of mean levels and variability in anxiety, depression, and sleep across COVID-19 groups

Figure 2 shows the distribution for anxiety, depression, and sleep mean scores per each group. Figure 3 represents the distribution of standard deviations of anxiety, depression and sleep per group.

**Fig 2.**
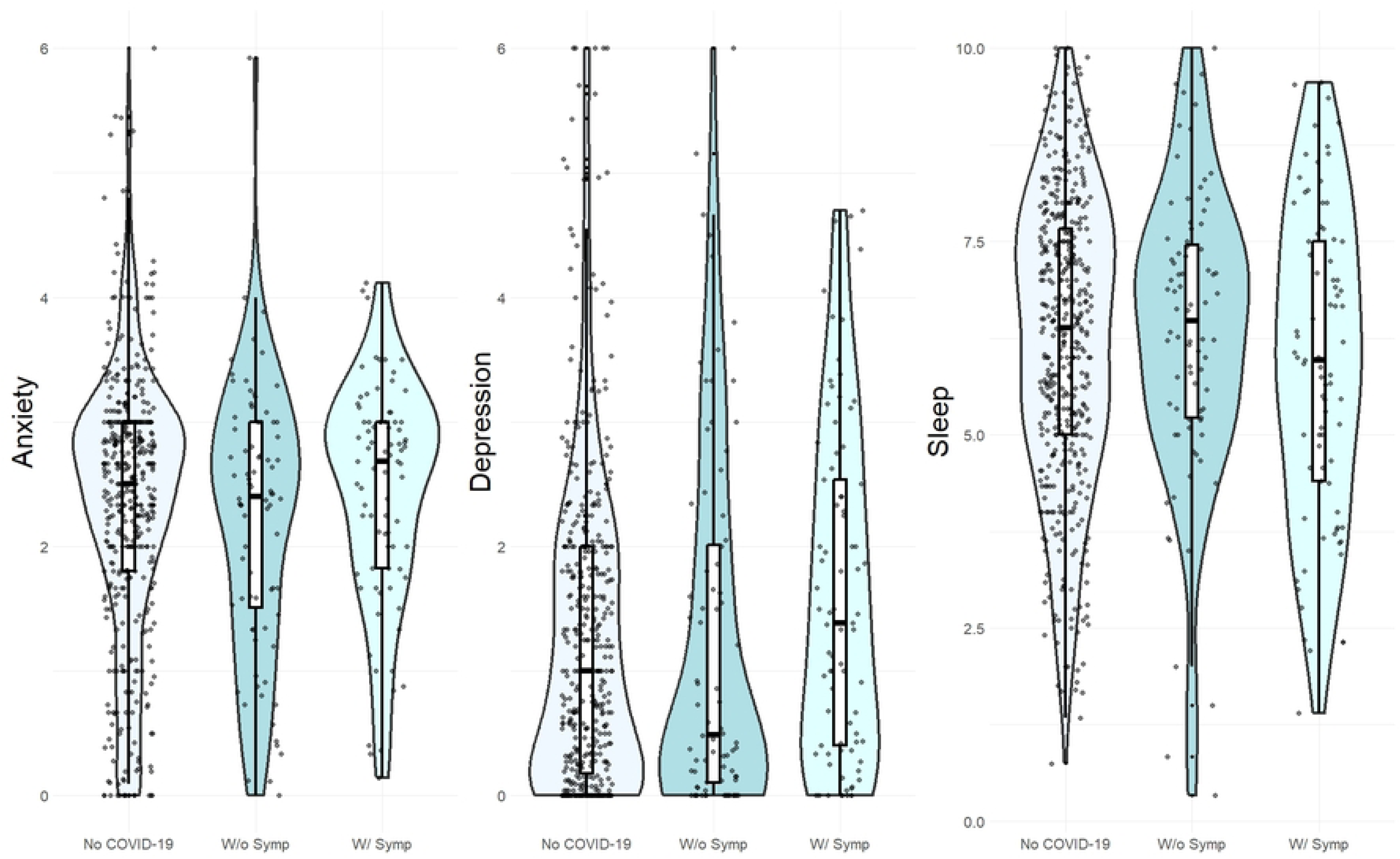
Violin plots for anxiety, depression and sleep quality mean scores n = 447 in the group without infection, n = 80 in the asymptomatic group, n = 81 in the group with COVID-19 symptoms

**Fig 3.**
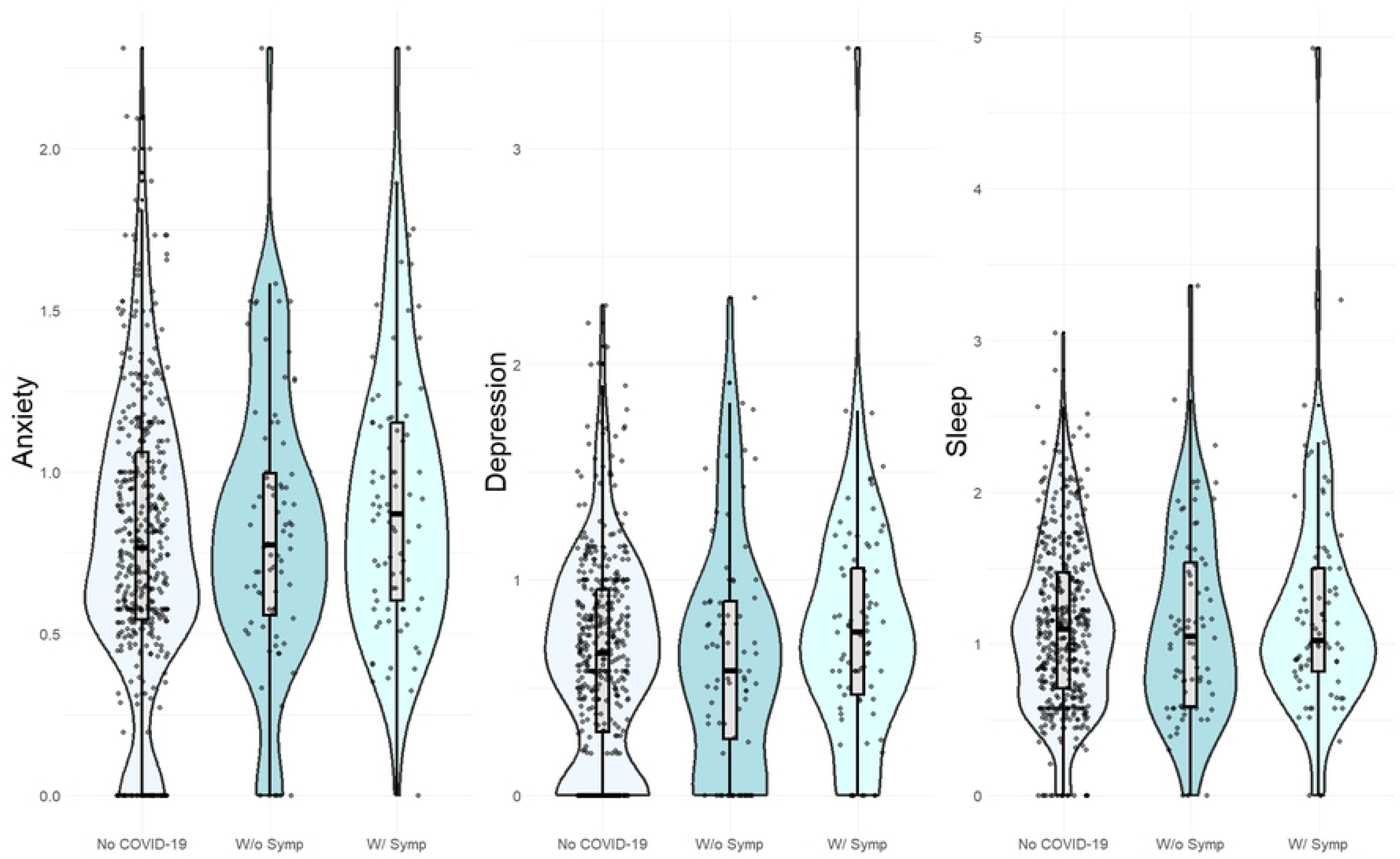
Violin plots for anxiety, depression and sleep quality standard deviation scores n = 447 in the group without infection, n = 80 in the asymptomatic group, n = 81 in the group with COVID-19 symptoms

Supplementary Tables 1 and 2 show the values of the multiple linear models for means and standard deviations, based on membership in the three groups, adjusting for baseline variables.

Examining the results of the multiple linear regressions for the mean scores of the three measures, no statistically significant differences were observed among the three groups (no COVID-19 diagnosis, symptomatic, and asymptomatic). The analysis revealed that women demonstrated significantly higher scores in anxiety and depression, as well as lower scores in sleep quality, indicating poorer outcomes across all three scales compared to men (p < 0.001).

Regarding the variability of the three scores, participants in the symptomatic group presented significantly higher variability for both anxiety and depression scores compared to the non-COVID-19 group (regression coefficients B = 0.112 [0.057, 0.167] and B = 0.124 [0.065, 0.183], respectively, with the confidence intervals calculated using the standard error). No significant difference was observed in outcome variability between the asymptomatic group and the non-COVID-19 group. Similarly, no significant effect of group membership on sleep variability was found.

To assess whether the time since SARS-CoV-2 infection influenced anxiety, depression, and sleep scores, ancillary analyses were performed using multiple linear models for the means and standard deviations of anxiety, depression, and sleep quality scores, specifically for infected individuals (see Supplementary Tables 3-4). The time since diagnosis did not significantly affect either the means or the standard deviations of the scores.

## Discussion

Our study assessed longitudinally anxiety, depression, and sleep quality among Spanish participants, focusing on differences between those who experienced COVID-19 symptoms, those who were asymptomatic, and those with no history of infection. While no significant differences were found in the mean levels of these mental health indicators and sleep quality across the three groups, our findings highlight an important aspect often overlooked in previous research: the greater variability in anxiety and depression scores observed among symptomatic individuals. This suggests that beyond mean levels, the experience of symptomatic COVID-19 may lead to increased instability in mental health over time, which has been linked to poorer long-term outcomes. In contrast, sleep quality did not exhibit similar variability, indicating a distinct trajectory for sleep disturbances in relation to COVID-19 symptoms. These findings contribute to ongoing discussions on the psychological burden of COVID-19, emphasizing the need to move beyond group-level comparisons of average scores and consider intra-individual fluctuations when evaluating mental health consequences.

The lack of significant difference between groups suggests that, when considering the overall mean scores across the three scales, there was no confirmed differences in mental health conditions among the three groups of participants, based on the presence or absence of COVID-19 symptoms. These results are in contradiction with the literature documenting the adverse mental health effects associated with SARS-CoV-2 infection [32–33]. Moreover, the analyses conducted by Tedjasukmana et al. and a systematic review and meta-analysis by Gui et al. confirm the worsening of sleep conditions following COVID-19 [34–35]. It is highly likely that the absence of a significant difference observed in our study is due to the time factor, as assessing mental health in the year after infection differs significantly from evaluating mental health during the acute phase of the disease at the time of infection [5].

Therefore, a key focus of the study was to account for variability within the data over time. A higher variability in anxiety and depression scores was observed in the symptomatic group compared to the non-COVID-19 group, indicating greater instability and worse mental health conditions, as greater variability is generally associated with poorer health outcomes [23–24]. Conversely, no differences were observed in the effect on sleep, which did not exhibit variability in the score in the three groups. Moreover, there were not significant differences between the asymptomatic and non-COVID-19 groups in terms of variability, highlighting that instability in mental health might predominantly be attributable to the manifestation of COVID-19 symptoms rather than the mere presence of the virus itself [36].

Gender differences emerged as a notable factor in our study, with women exhibiting higher levels of depression and anxiety, as well as poorer sleep quality compared to men. This finding is consistent with existing literature, which indicates that women are at a higher risk for stress and anxiety [37], and it aligns with research showing that women experienced significant sleep disturbances during the COVID-19 pandemic, which were closely associated with increased levels of anxiety and depression [38]. Furthermore, our analysis suggests that women may experience a greater risk of post-COVID-19 condition, adversely affecting their quality of life and leading to heightened long-term concerns related to COVID-19 [39]. Further, we did not find any significant associations between COVID-19 vaccination status and scores for anxiety, depression, or sleep quality. This contrasts with the study by Nguyen, which suggests that COVID-19 vaccination positively influences mental health by reducing symptoms of anxiety and depression, as the sense of protection associated with high vaccination rates may enhance individuals’ psychological well-being [40]. A relationship was also observed between vaccination, antibody levels, and mental illness, with individuals with mental disorders tending to exhibit lower antibody levels in response to vaccination compared to those without such conditions, two years after the outbreak and the vaccination phase [41].

Given the robust correlation established between COVID-19 symptoms and the exacerbation of mental health disorders, it is crucial to prioritize the implementation of preventive mental health programs aimed at enhancing overall quality of life of infected patients. Psychological-behavioural intervention (PBI) programs, as suggested by Kong et al., represent a promising avenue for addressing the emerging mental health challenges associated with the pandemic [42]. When planning such interventions, it is important not only to focus on addressing the mean levels of mental health but also to consider the variability in mental health outcomes. These interventions could help mitigate psychological distress experienced by symptomatic individuals, thereby fostering resilience and improving mental health outcomes in the face of ongoing public health challenges.

### Strengths and limitations

The main strength of this study lies in its longitudinal design, which allows for the assessment of changes in mental health over time, providing a more accurate understanding of the long-term effects of COVID-19 and characterizing variability within each subject over time, beyond mean levels.

The study also had limitations. We focused on anxiety, depression, and subjective sleep quality, whereas other mental health outcomes, such as mood and affect, were not examined. Secondly, while our analysis of COVID-19 symptoms enabled comparisons between participants based on the presence or absence of these symptoms, it did not allow for an exploration of the varying severities of COVID-19. This limitation was particularly significant due to the small number of participants who experienced severe symptoms, which constrained our ability to draw comprehensive conclusions regarding the impact of symptom severity on mental health and sleep outcomes. Thirdly, the analyses were conducted on a sample that does not include young people, a particularly vulnerable group that experienced a significant increase in the prevalence of depression and anxiety following the initial outbreak.

## Conclusions

The primary findings of this study suggest that individuals exhibiting COVID-19 symptoms experience significantly greater variability in anxiety and depression scores over time following infection, indicating that their mental health conditions are marked by increased instability and fluctuations, which, according to the literature, are associated with poorer health outcomes. In contrast, the data suggest that sleep quality remains largely unaffected by the presence of COVID-19 symptoms at baseline, highlighting a potential dissociation between COVID-19 symptomatology and sleep disturbances over time. These results underscore the necessity for targeted mental health interventions that address the specific challenges faced by symptomatic individuals, while also emphasizing the need to consider not only the mean levels of mental health but also the variability over time when providing and evaluating interventions. Moreover, it is crucial to prioritize women’s well-being, as their quality of life has declined post-COVID-19.

## Statements & Declarations

### Data Availability

The R code and the datasets generated during and analysed during the current study are available in the open science framework page of the project, https://doi.org/10.17605/OSF.IO/QCR6J.

### Author contributions

Marco Viola: Conceptualization (equal); Material preparation; Methodology (equal); Formal analysis; Writing – original draft; Writing – review and editing (equal).

Guillaume Chevance: Conceptualization (equal); Supervision (equal); Project Administration (equal); Methodology (equal); Writing – review and editing (equal).

Manolis Kogevinas: Supervision (equal); Project Administration (equal); Methodology (supporting); Writing – review and editing (supporting).

Rosalba Rosato: Methodology (supporting); Writing – review and editing (supporting). Gemma Castaño-Vinyals: Writing – review and editing (supporting).

Rafael de Cid: Writing – review and editing (supporting). All authors read and approved the final manuscript.

### Ethical standards

The authors assert that all procedures contributing to this work comply with the ethical standards of the relevant national and institutional committees on human experimentation and with the Helsinki Declaration of 1975, as revised in 2008.

Approval was granted by The Parc de Salut Mar Ethics Committee (CEIm-PS MAR, number 2020/9307/I).

### Supporting information

S1 Supplementary Material

